# Recapitulation of Cellular Senescence, Inflammation, and Fibrosis in Human Kidney-Derived Tubuloids by Repeated Cisplatin Treatment: A Novel Pathophysiological Model for Sensing Nephrotoxicity and Drug Screening

**DOI:** 10.1101/2024.03.17.24304404

**Authors:** Yuki Nakao, Makiko Mori, Yuta Sekiguchi, Iori Morita, Ryota Shindoh, Shintaro Mandai, Tamami Fujiki, Hiroaki Kikuchi, Fumiaki Ando, Koichiro Susa, Takayasu Mori, Yuma Waseda, Soichiro Yoshida, Yasuhisa Fujii, Eisei Sohara, Shinichi Uchida, Yutaro Mori

**Affiliations:** Department of Nephrology, Graduate School of Medical and Dental Sciences, Institute of Science Tokyo, Tokyo 113-8510, Japan; Department of Urology, Graduate School of Medical and Dental Sciences, Institute of Science Tokyo, Tokyo 113-8510, Japan

**Keywords:** organoid, cisplatin nephrotoxicity, fibrosis, chronic kidney disease (CKD)

## Abstract

In the pursuit of pathophysiological models for assessing renal drug response, the development of kidney organoids derived from human pluripotent stem cells represents a significant step forward. However, recapitulating aging/senescence-associated pathophysiology remains challenging. Here, we present an innovative approach to generate epithelial-like structures known as “tubuloid” using primary human renal proximal tubular epithelial cells (hRPTECs) cultured from human resected kidneys, as a refined alternative. We evaluated the efficacy of tubuloids using cisplatin treatment. Tubuloids showed highly differentiated structures. Exposure to cisplatin increased γH2AX, Kidney Injury Molecule-1 (KIM-1) and Cleaved Caspase-3, markers for DNA damage response, epithelial damage and apoptosis respectively. Repeated cisplatin administration resulted in upregulation of the cellular senescence marker p16, p21 and SA-β-Gal. Additionally, increased secretion of inflammatory cytokines, indicating the induction of a senescence-associated secretory phenotype (SASP) were induced Supernatant collected from cisplatin-treated tubuloids induced myofibroblast activation, indicating the onset of renal fibrosis. We successfully established a tubuloid-based model of cisplatin-induced kidney injury using hRPTECs. Tubuloids can replicate cellular senescence, SASP, and fibrosis, which can recapitulate the phenotypes of chronic kidney disease (CKD). Furthermore, tubuloids provide a novel platform for studying the response of renal epithelial cells to toxins and therapeutics and offer innovative strategy for drug screening in a human-based fashion.

**Translational Statement:** Recapitulating aging/senescence-associated pathophysiological reaction in kidney organoids remains challenging. Our study reveals that tubuloids could be novel candidate for chronic kidney disease (CKD) model.

## Introduction

Chronic kidney disease (CKD) is an irreversible, progressive condition characterized by the gradual sclerosis of glomeruli, accompanied by tubular atrophy, interstitial fibrosis, and infiltration of inflammatory cells [1]. Despite its significant impact on global health, the underlying pathogenic mechanisms of CKD are poorly understood, and a definitive cure has yet to be discovered.

In recent years, a shared mechanism for inflammation and fibrosis in CKD has emerged. It is proposed that the initial trigger is DNA damage in tubular epithelial cells, which initiates a cascade involving DNA damage response, cell cycle arrest [2], cellular senescence [3, 4], and the subsequent development of the senescence-associated secretory phenotype (SASP) [5]. To investigate these processes, various models have been created, including cell and animal models, as well as kidney organoids derived from human induced pluripotent stem cells (iPSCs) [6] or human embryonic stem cells (ESCs). These models show promise in understanding the pathogenesis of CKD and investigating potential therapeutic interventions [7]. Nonetheless, one critical limitation of existing models is their inability to fully capture the aging process, particularly the phenomenon of cellular senescence, which manifests prominently in humans with a lifespan greater than 80 years. Experimental animals such as mice, with a lifespan of only two years, do not fully replicate the range of age-related diseases associated with cellular senescence found in humans.

Furthermore, despite progress, kidney organoids derived from iPSCs or ESCs have primarily focused on replicating fetal kidney development stages, falling far short of accurately mimicking age-related diseases such as CKD.

In this context, “tubuloids,” a three-dimensional structure resembling renal tubules, were first reported in 2019. These tubuloids come from adult primary human renal proximal tubular epithelial cells (hRPTECs) [8]. Schutgens et al. pioneered the culture of primary hRPTECs derived from adult human kidneys and urine, successfully producing tubuloids capable of replicating viral infections in proximal tubules and hereditary diseases [9]. Building on this work, we have further refined the tubuloid generation protocol with human resected kidneys [10]. Our research has focused on understanding the response of tubuloids to a nephrotoxicant and a potential therapeutic agent. Despite these advances, developing a comprehensive pathophysiological model for aging or senescence-associated diseases using tubuloids remains a promising area of research.

Cisplatin, a commonly used therapeutic agent for various malignancies, including lung cancer, has a well-known side effect known as cisplatin-induced nephropathy [11, 12]. This condition is defined by the induction of DNA damage in the renal proximal tubules [13]. The cellular response to this damage includes cell cycle arrest during the repair phase following cisplatin-induced injury, which eventually leads to cellular senescence and SASP. These processes are thought to be the final detrimental mechanisms that underpin the progression from acute kidney injury (AKI) to CKD [14, 15]. Thus, modeling cisplatin-induced nephropathy is not only useful for studying nephrotoxicity but also has the potential to mimic the progression of CKD.

The primary goal of this study was to assess the disease-modeling potential of tubuloids by exposing them to cisplatin to reproduce cisplatin-induced nephropathy. Furthermore, we wanted to investigate whether tubuloids could replicate cellular senescence and its associated responses beyond the immediate effects of cisplatin nephrotoxicity to simulate the transition from AKI to CKD, or even model CKD itself.

## Methods

### Cell culture experiment

Human kidney samples were collected from patients undergoing clinically indicated nephrectomy at the Tokyo Medical and Dental University Hospital in Tokyo, Japan. The protocol was approved by the Institutional Review Board of the Ethics Committee of Tokyo Medical and Dental University (M2022-005). Human renal proximal tubular epithelial cells (hRPTECs) were isolated from unaffected areas of kidneys removed during nephrectomy procedures performed on patients with renal cell carcinoma or urinary tract malignancies by modifying a previously established protocol[16]. Briefly, the isolation procedure involved mincing the human renal cortex and then digesting it in a solution of Collagenase type II (1.0 mg/mL) (Worthington Biochemical, NJ, USA). The enzyme reaction was terminated with fetal bovine serum (FBS), and the samples were resuspended in hRPTECs culture medium containing DMEM/F-12 (Nacalai Tesque, Kyoto, Japan), with BSA (Nacalai Tesque), Antibiotic– Antimycotic (ThermoFisher Scientific, MA, USA), hydrocortisone (ThermoFisher Scientific, MA, USA), ITS liquid media supplement (Sigma-Aldrich, MO, USA), and human recombinant epidermal growth factor (EGF) (ThermoFisher Scientific, MA, USA). The epithelial cells were cultured for 7-10 days before being used in experiments.

To make mouse primary kidney fibroblasts (MPKFs), a 12-week old male C57BL/6J mouse was sacrificed. MPKFs were isolated by mincing the mouse kidneys and digesting them in a solution of Collagenase type II (1.0 mg/mL). The enzyme reaction was stopped with FBS, and the samples were resuspended in DMEM with 10% FBS, 5 ng/ml FGF-basic (ThermoFisher Scientific), and Antibiotic-Antimycotic. The MPKFs were used after 3 to 5 passages. All cells were maintained in a CO_2_ incubator (5% CO_2_) at 37°C to ensure optimal growth conditions. All animal studies were performed in accordance with the guidelines for animal research of Tokyo Medical and Dental University. The study protocol was approved by the Animal Care and Use Committee of Tokyo Medical and Dental University (approval no. A2023-109C).

### Human Renal Tubuloids Culture

hRPTECs were seeded onto ultralow attachment plates at a density of 5.0 × 10^5^ cells/well using Advanced RPMI 1640 medium (ThermoFisher Scientific), supplemented with 5% FBS. After a 2-day incubation period, Matrigel (Corning, NY, USA) was added to stimulate tubuloid formation. The following day, the culture medium was changed to Advanced RPMI 1640 medium with 5% FBS, EGF, FGF2, and HGF (tubuloids media). Media was changed once or twice weekly to maintain optimal cell growth conditions. Tubuloids were usually ready for experimentation within 1–2 weeks of starting the culture.

### Fibrosis bioassay

The tubuloids were treated with or without cisplatin on a repeated basis. Following a 48-hour exposure period, the media was replaced with serum-free DMEM to facilitate conditioning. After an additional 48 hours of incubation, the culture supernatants were collected as conditioned media.

MPKFs were uniformly plated at a density of 2.0 × 10^4^ cells/well in an 8-well chamber slide (Nunc Lab-Tek Chamber Slide system) (ThermoFisher Scientific) with 10% FBS-DMEM. The next day, the conditioned media harvested from the tubuloids was added to MPKFs and incubated for 48 hours. The MPKFs on the 8-well chamber slide were then washed twice with PBS and immunostained as described below.

### Immunofluorescence staining

The tubuloids were fixed with 4% paraformaldehyde (PFA) in PBS and permeabilized with 0.1% Triton X-100 in PBS. The tubuloids were then blocked with 3% BSA in PBS and incubated with primary antibodies for 1 hour. Tubuloids were washed thoroughly with PBS and then incubated with secondary antibodies for 30 minutes. After another round of PBS washing, the tubuloids were placed on glass slides. The slides were mounted with Prolong Glass Antifade Mountant with NucBlue Stain (ThermoFisher Scientific) and cover-slipped for fluorescence microscopy.

The following primary antibodies were used: rabbit anti-LRP2 / Megalin antibody (1:100) (ab236244, Abcam, MA, USA), rabbit anti-γH2AX antibody (1:200) (#9718, Cell Signaling Technology, MA, USA), mouse anti-Na^+^/K^+^-ATPase antibody (1:200) (05-369, Sigma-Aldrich), goat anti-KIM-1 antibody (1:200) (AF1750, R&D systems, MN, USA), rabbit anti-caspase-3 antibody (1:200) (#9664, Cell Signaling Technology), rabbit anti-Vimentin antibody (1:200) (10366-1-AP, Proteintech, IL, USA), rabbit anti-p16-INK4A polyclonal antibody (1:200) (10883-1-AP, Proteintech), rabbit anti-IL-1β polyclonal antibody (1:200) (bs-0812R, Bioss, MA, USA), rabbit anti-Collagen type 1 antibody, rabbit anti-αSMA antibody conjugated with Cy3 (1:400) (C6198, Sigma-Aldrich) and rabbit anti-collagen type 1 antibody (1:250) (600-406-103, rockland) were used. To stain the brush border of the proximal tubular epithelium, LTL, Biotinylated (B-1325-2, Vector Laboratories, CA, USA) was used instead of primary antibodies (1:200). Streptavidin, Alexa Fluor 633 conjugate (S21375, ThermoFisher Scientific) was then applied (1:500).

For the immunofluorescence staining of fibrosis bioassay, MPKFs plated on an 8-well chamber slide were washed before being fixed with 4% PFA-PBS. The cells were permeabilized with 0.1% Triton X-100 in PBS and then blocked with 3% BSA.

All images were captured using either standard or confocal microscopy (Eclipse Ti2 from Nikon), with the NIS-Elements Advanced Research microscope system serving as the operating system. Quantification of stained area was done using ImageJ Fiji (https://imagej.net/software/fiji/downloads).

### Quantitative reverse transcription-polymerase chain reaction (qRT-PCR)

The qRT-PCR analysis of human IL-1β, IL-6, and 18S mRNA followed established protocols. The real-time PCR detection system utilized was the Thermal Cycler Dice Real Time System Lite TP700 (Takara Bio, Shiga, Japan) along with TB green Premix Ex Taq II (Takara Bio, Shiga, Japan) [17]. The total RNA was extracted from frozen kidney tubuloids stored at −80°C using Sepasol-RNAI Super G (Nacalai Tesque, Kyoto, Japan). cDNA was then synthesized from the isolated total RNA using ReverTra Ace (TOYOBO, Tokyo, Japan) [18]. The generated cDNA was then amplified through 40 PCR cycles, with denaturation at 95°C for 5 seconds and annealing/extension at 60°C for 30 seconds.

The nucleotide sequences of the primers utilized for qRT-PCR analysis are as follows. human 18S.

Forward:5′-GCAGAATCCACGCCAGTACAAG-3′ Reverse:5′-GCTTGTTGTCCAGACCATTGGC-3′

human IL-1β.

Forward: 5′- CCACAGACCTTCCAGGAGAATG -3′

Reverse: 5′- GTGCAGTTCAGTGATCGTACAGG -3′

human IL-6.

Forward 5′- AGACAGCCACTCACCTCTTCAG -3′

Reverse: 5′- TTCTGCCAGTGCCTCTTTGCTG -3′

### Western blot analysis

The tubuloids were lysed, and the protein was purified. Protein bands were visualized using Western Blue (Promega, WI, USA). Rabbit anti–ERK2 (p44/42 MAPK) antibody (9102S, Cell Signaling Technology) (1:200) was used as a loading control. Primary antibodies were also used, such as rabbit anti-p16-INK4A polyclonal antibody (1:200) (Proteintech), mouse anti-p21 antibody (1:100) (sc-6247, Santa Cruz Biotechnology, TX, USA), rabbit anti-KIM-1 antibody (1:400) (PA5-20244, Thermo Fisher Scientific, MA, USA), rabbit anti-caspase-3 antibody (1:200) (Cell Signaling Technology) and rabbit anti-Vimentin antibody (1:1000) (#5741, Cell Signaling Technology, MA, USA). Densitometry was performed by using Image J Fiji (https://imagej.net/software/fiji/downloads).

### Senescence-associated-β-galactosidase (SA-β-GAL) staining

The tubuloids were stained by using Cellular Senescence Detection Kit (SA-β-Gal Staining) (CBL, CA, USA) according to directions of the manufacturer. Four wells of organoids cultured in a 24-well plate were used for each experimental condition, and images were captured to encompass all areas of the wells.

### Flowcytometry

Tubuloids were digested in using 2.5g/l-Trypsin/1mmol/l-EDTA Solution, with Phenol Red (Nacalai Tesque Inc., Kyoto, Japan) at 37 degree for 20 minutes. Single cells obtained from tubuloids were fixed with 4% PFA-PBS with 5% FBS, permeabilized with 0.1% Triton X-100-PBS, and stained with primary antibodies for 1 hour and with secondary antibodies for 45 minutes. Rabbit anti-LRP2 / Megalin antibody (1:250) (Abcam) and rabbit anti-caspase-3 antibody (1:250) (Cell Signaling Technology) were used for primary antibodies. Cells were analyzed by BD FACSLyric (BD Biosciences, MA, USA). Data were analyzed by FlowJo (BD Biosciences, MA, USA).

### Quantification and Statistical Analysis

Figure Legends specify the number of samples assayed in each experiment. To determine significant differences between groups, a one-way analysis of variance was used. A p-value of less than 0.05 was considered statistically significant. All statistical analyses were performed with GraphPad Prism by GraphPad Software Inc. (San Diego, CA, USA).

## Results

### Primary human renal proximal tubular epithelial cells (hRPTECs) form highly polarized structure

To form tubuloids, human renal proximal tubular epithelial cells (hRPTECs) were isolated from the cortex of human kidneys, embedded in a basement membrane gel, and cultured with various growth factors **(Figure 1A)**. Using precise techniques that included finely mincing the cortex portion of the human kidneys and selective culturing of epithelial cells in a serum-free medium supplemented with epidermal growth factor (EGF), primary cultured cells, primarily proximal tubular epithelial cells, were successfully established **(Figure 1B and 1C)**.

**Figure 1.**
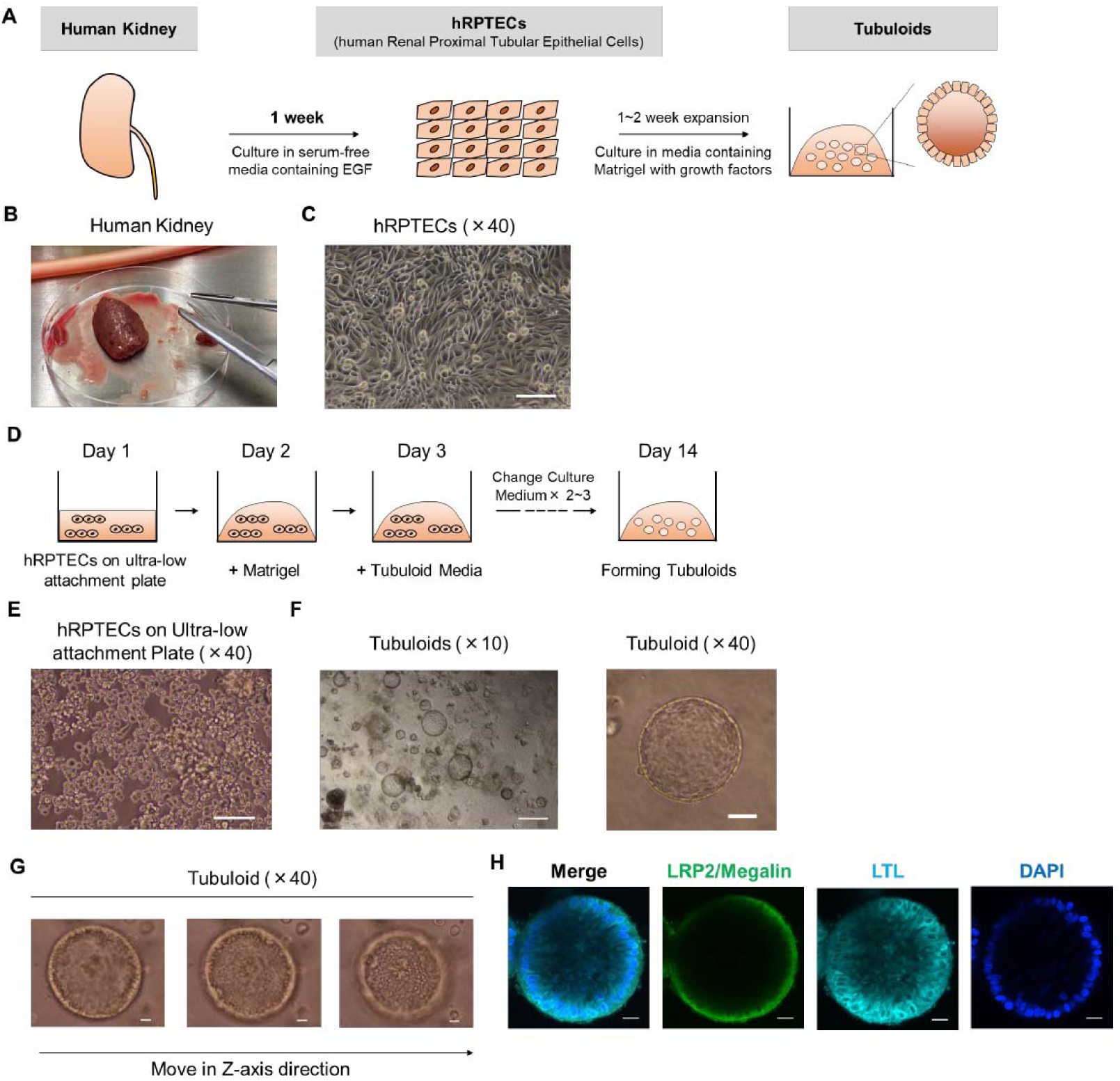
Formation of highly polarized tubuloid structures from primary human renal proximal tubular epithelial cells (hRPTECs). (A) Schematic overview of the establishment of tubuloids from hRPTECs isolated from adult human kidney. (B) Image of the human resected kidney utilized during the establishment of hRPTECs. (C) Representative image showing hRPTECs. Scale bars: 50 µm. (D) Two-week protocol for generating tubuloids from 2D-cultured hRPTECs. (E) Representative image demonstrating the seeding of hRPTECs on an ultralow attachment plate. Scale bars: 25 µm. (F) Representative images of tubuloids at different magnifications (×10, ×40). Scale bars: 100 µm, 25 µm. (G) Representative image of tubuloids at ×40 magnification, showing multiple layers along the Z axis. Scale bars: 25 µm. (H) Immunostaining of makers for renal proximal tubular epithelial cells (LRP2/Megalin and LTL). Scale bars: 25 µm.

Tubuloids were then formed over approximately a two-week protocol **(Figure 1D).** Specifically, hRPTECs were seeded on an ultralow attachment plate **(Figure 1E),** followed by the addition of Matrigel and media containing fetal bovine serum, EGF, fibroblast growth factor 2 (FGF2), and hepatocyte growth factor (HGF). Multiple tubuloids were formed in a single well, with nearly uniform sizes. The epithelial cells formed tubular-like structures that encircled the basement membrane gel **(Figure 1F)**. The three-dimensional architecture was confirmed by manipulating the microscope along the Z-axis direction **(Figure 1G)**. Notably, the cells formed a monolayer structure similar to tubules or renal cysts, rather than a spheroid-like structure. Furthermore, tubuloids expressed differentiation markers associated with proximal tubular epithelial cells, such as lotus tetragonolobus lectin (LTL) and LDL Receptor Related Protein 2 (LRP2/Megalin), confirming their highly differentiated and polarized structure **(Figure 1H)**[19].

### Cisplatin Exposure Induces Tubular Structure Collapse and Elicits Acute DNA Damage Response (DDR) in Tubuloids

To test the sensitivity of tubuloids against a nephrotoxicant, we exposed them to cisplatin. Cisplatin causes cytotoxicity by entering proximal tubular epithelial cells primarily through Organic Cation Transporter 2 (OCT2) [20], resulting in direct DNA damage **(Figure 2A)**. Consistent with previous reports highlighting the activation of DNA damage response (DDR) pathways, we found significant upregulation of γH2AX, a marker of DNA double-strand breaks, in both *in vivo* and *in vitro* [21]. Initially, tubuloids were treated with cisplatin at three concentrations of 0.2, 2.0, and 20.0 µg/mL to determine dose-dependent responses. Notably, high-dose cisplatin treatment collapsed the three-dimensional tubuloid architecture, **(Figure 2B)**, demonstrating the extent of DNA damage caused by cisplatin exposure **(Figure 2C-E).** Furthermore, immunostaining analyses revealed a significant reduction in the expression levels of differentiation markers such as LRP2 and Na+/K+-ATPase after cisplatin treatment. This downregulation indicates a concomitant process of cellular dedifferentiation, possibly as an adaptive response to cytotoxic insult **(Figures 2F-K).**

**Figure 2.**
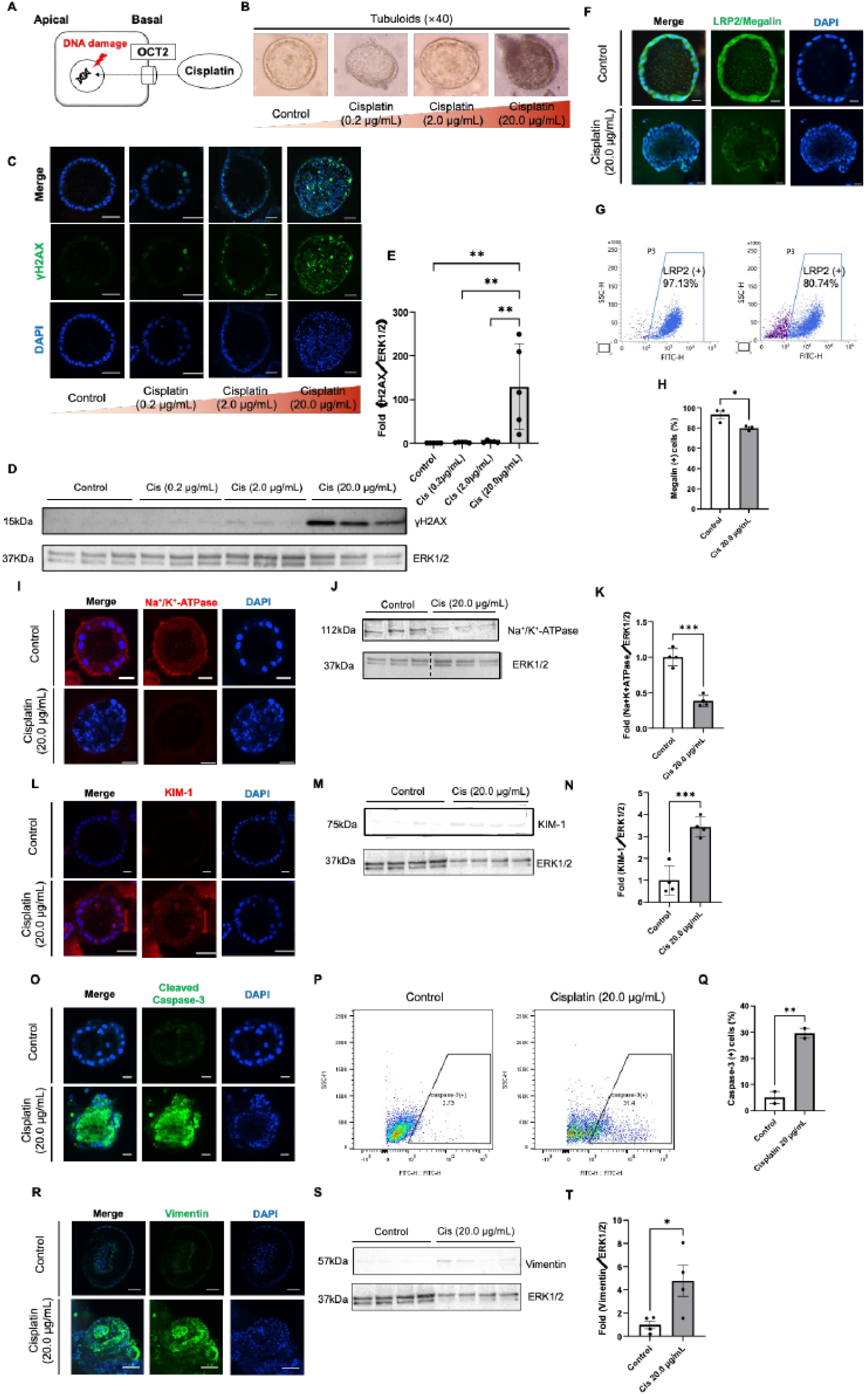
Effects of cisplatin treatment on tubuloid structure, DNA damage response, differentiation loss, cell death, and epithelial-mesenchymal transition. (A) Schematic representation of the pathway by which cisplatin induces DNA damage in proximal tubular epithelial cells. (B) Representative images of tubuloids exposed to cisplatin (0.2, 2.0, 20.0 µg/mL) or control. (C) Immunostaining of marker for DNA damage (γH2AX) in tubuloids exposed to cisplatin (0.2, 2.0, 20.0 µg/mL) or control. Scale bars: 100 µm. (D) Western blotting of γH2AX in tubuloids exposed to cisplatin (0.2, 2.0, 20.0 µg/mL) or control. (E) Statistical analysis of γH2AX expression (n = 4). (F) Immunostaining of a differentiation marker for epithelial cells, LRP2/Megalin in tubuloids exposed to cisplatin (20.0 µg/mL) or control. Scale bars: 25 µm. (G) Representative plots of flowcytometry measuring LRP2/Megalin expression in tubuloids exposed to cisplatin (20.0 µg/mL) or control. (H) Statistical analysis of LRP2/Megalin expression (n = 3). (I) Immunostaining of a differentiation marker for epithelial cells, Na^+^/K^+^-ATPase in tubuloids exposed to cisplatin (20.0 µg/mL) or control. Scale bars: 25 µm. (J) Western blotting of Na^+^/K^+^-ATPase in tubuloids exposed to cisplatin (20.0 µg/mL) or control. (K) Statistical analysis of LRP2/Megalin expression (n = 4). (L) Immunostaining of an epithelial injury marker, Kidney Injury Molecule-1 (KIM-1) in tubuloids exposed to cisplatin (20.0 µg/mL) or control. Scale bars: 25 µm. (M) Western blotting of KIM-1 in tubuloids exposed to cisplatin (20.0 µg/mL) or control. (N) Statistical analysis of KIM-1 expression (n = 4). (O) Immunostaining of an apoptosis marker, Cleaved Caspase-3, in tubuloids exposed to cisplatin (20.0 µg/mL) or control. Scale bars: 25 µm. (P) Representative plots of flowcytometry measuring Cleaved Caspase-3 expression in tubuloids exposed to cisplatin (20.0 µg/mL) or control. (Q) Statistical analysis of Cleaved Caspase-3 expression (n = 4). (R) Immunostaining of intermediate filament, Vimentin, in tubuloids exposed to cisplatin (20.0 µg/mL) or control. Scale bars: 100 µm. (S) Western blotting of Vimentin in tubuloids exposed to cisplatin (20 µg/mL) or control. (T) Statistical analysis of Vimentin expression (n = 4). All data are presented as mean ± standard error (SE). Significance is presented by *p < 0.05, **p < 0.01, ***p < 0.001.

Additionally, there was an increase in the expression of a renal injury marker, Kidney Injury Molecule-1 (KIM-1) [19], and an apoptosis marker, Cleaved Caspase-3 **(Figure 2L-Q)**, which was consistent with previous findings [20]. These findings further support the sensitivity of tubuloids to cisplatin-induced injury and validate their use as a model system for studying acute responses, including DDR.

Interestingly, we found that high concentrations of cisplatin increased the expression of Vimentin, an intermediate filament specific to mesenchymal cells **(Figure 2R-T).** This suggests that proximal tubular epithelial cells may undergo epithelial-mesenchymal transition (EMT) as part of a tissue repair mechanism [22, 23]. The activation of Vimentin following DDR has been reported previously [24], and our study confirms this sequential response in cisplatin-treated tubuloids, elucidating the molecular dynamics underlying cisplatin-induced nephrotoxicity.

### Tubuloids show cellular senescence and release inflammatory cytokines following repeated cisplatin treatment

To assess the potential for replicating chronic responses, tubuloids were subjected to prolonged treatment of cisplatin. Initially, cisplatin was administered at concentrations of 0.2 and 2.0 µg/mL. The media was then replaced three times to reduce residual cisplatin, and standard culture conditions were applied for one day, completing one cycle. This was considered one cycle, and the next day, cisplatin was given again at similar concentrations **(Figure 3A)**. The cycle was repeated five times. Although treatment with 20.0 µg/mL cisplatin caused significant disruption of tubuloids and cell death **(Figure 2)**, this concentration was excluded from the experimental setup to focus on long-term chronic responses.

**Figure. 3.**
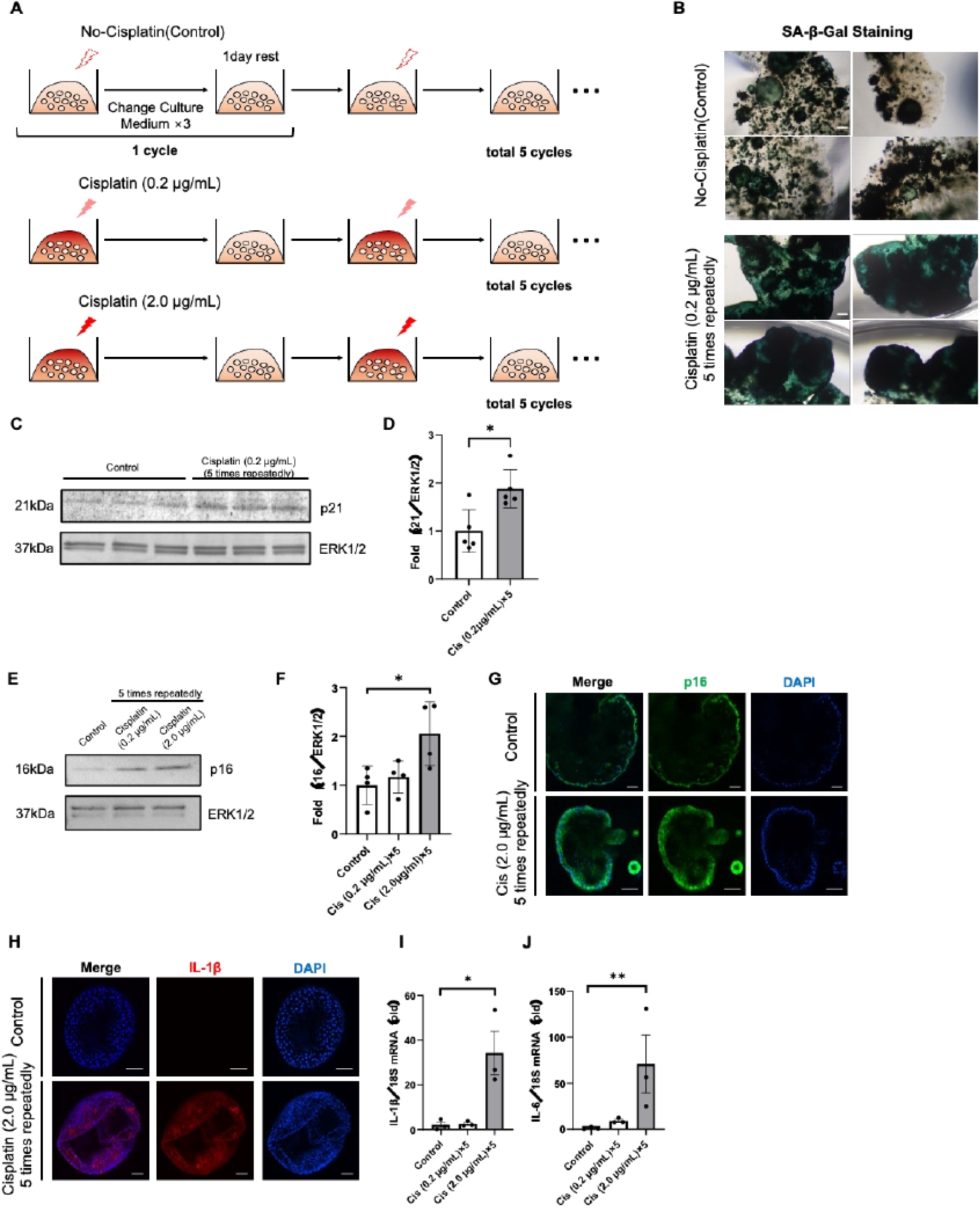
Tubuloids exhibit cellular senescence and release inflammatory cytokines following repeated cisplatin treatment. (A) Experimental design for repeated cisplatin injury. (B) Representative images of SA-β-GAL staining of tubuloids exposed to cisplatin (0.2 µg/mL) 5 times repeatedly or control. Scale bars: 100 µm. (C) Western blotting of p21 in tubuloids exposed to cisplatin (0.2 µg/mL) 5 times repeatedly or control. (D) Statistical analysis of p21 expression (n = 4). (E) Western blotting of p16 in tubuloids exposed to cisplatin (0.2, 2.0 µg/mL) 5 times repeatedly or control. (F) Statistical analysis of p16 expression (n = 4). (G) Immunostaining of p16 in tubuloids exposed to cisplatin (2.0 µg/mL) 5 times repeatedly or control. Scale bars: 100 µm. (H) Immunostaining of an inflammatory cytokine, IL-1β, in tubuloids exposed to cisplatin (2.0 µg/mL) 5 times repeatedly or control. Scale bars: 100 µm. (I, J) Quantitative PCR of IL-1β (I) and IL-6 (J) in tubuloids exposed to cisplatin (0.2, 2.0 µg/mL) 5 times repeatedly or control (n = 3). Statical analysis was applied to delta-delta Ct value, not fold increase. All data are presented as mean ± standard error (SE). Significance is presented by *p < 0.05, **p < 0.01.

In the repeated cisplatin administration model, we observed the expression of p16, p21 and SA-β Gal, which are well-established **(Figure 3B-G)** and consistent with previous findings [25–27]. Additionally, we confirmed an increased expression of the inflammatory cytokine IL-1β at a concentration of 2.0 µg/mL through immunofluorescence analyses **(Figure 3H)**. Quantitative PCR analysis revealed a significant upregulation in mRNA levels of both IL-1β and IL-6, ranging from 50 to 100-fold compared to controls, in the group subjected to repeated administration of 2.0 µg/mL cisplatin (*P = 0.0167*; IL-1β and *P = 0.011*; IL-6) **(Figure 3I and J**). These findings reveal that repetitive DNA damage induced by cisplatin caused cellular senescence, which resulted in the acquisition of an SASP.

### Secretions released from cisplatin-treated tubuloids strongly promote myofibroblast differentiation *in vitro*

We hypothesized that prolonged exposure of tubuloids to high concentrations of cisplatin would promote myofibroblast activation, potentially contributing to tissue fibrosis via the secretion of paracrine factors from tubuloids, as previously reported *in vivo* [28]. To test this hypothesis, we used a fibrosis bioassay, as described previously [10] **(Figure 4A)**. Initially, tubuloids were divided into three groups: one receiving no cisplatin, one receiving 0.2 µg/mL cisplatin, and another receiving 2.0 µg/mL cisplatin. Each group received repeated treatments. Following cisplatin administrations, the medium was changed three times to remove residual cisplatin, and the tubuloids were cultured for an additional 2 days in serum-free conditions to produce conditioned media (CM) containing secreted paracrine factors. Subsequently, the collected CM was administered to fibroblasts derived from the mouse renal cortex **(Figure 4B)**. The activation of myofibroblasts was assessed by evaluating the expression of α-smooth muscle actin (α-SMA) and Collagen type I[29].

**Figure 4.**
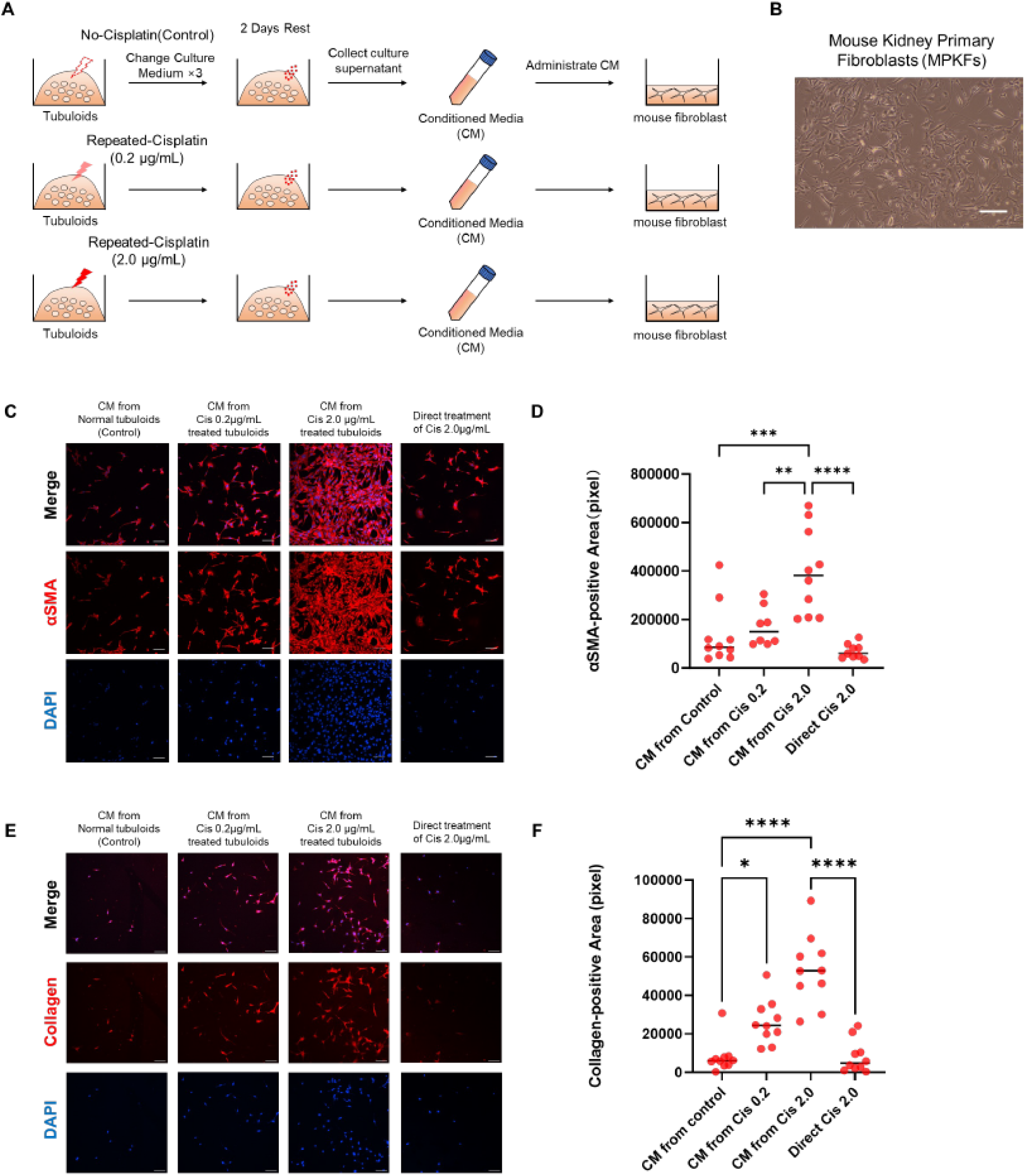
Secretion released from cisplatin-treated tubuloids strongly promotes fibrosis *in vitro*. (A) Experimental design for fibrosis bioassay. (B) A representative image of Mouse Primary Kidney Fibroblasts (MPKFs). Scale bar: 25 µm. (C) Representative images of fibrosis bioassay stained with anti-α-SMA antibody on MPKFs cultured with Conditioned Media (CM) harvested from tubuloids exposed to cisplatin (0.2, 2.0 µg/mL) or control, or directly with cisplatin-containing media. Scale bars: 100 µm. (D) Quantification of αSMA-positive area in MPKFs cultured with CM or directly with cisplatin-containing media measured by Image J (*n* = 8-10 in each condition). (E) Representative images of fibrosis bioassay stained with anti-Collagen type I antibody on MPKFs cultured with Conditioned Media (CM) harvested from tubuloids exposed to cisplatin (0.2, 2.0 µg/mL) or control, or directly with cisplatin-containing media. Scale bars: 100 µm. (F) Quantification of Collagen type I-positive area in MPKFs cultured with CM or directly with cisplatin-containing media measured by Image J (*n* = 8-10 in each condition). All data are presented as mean ± standard error (SE). Significance is presented by *p < 0.05, **p < 0.01, ***p < 0.001, ****p <0.0001.

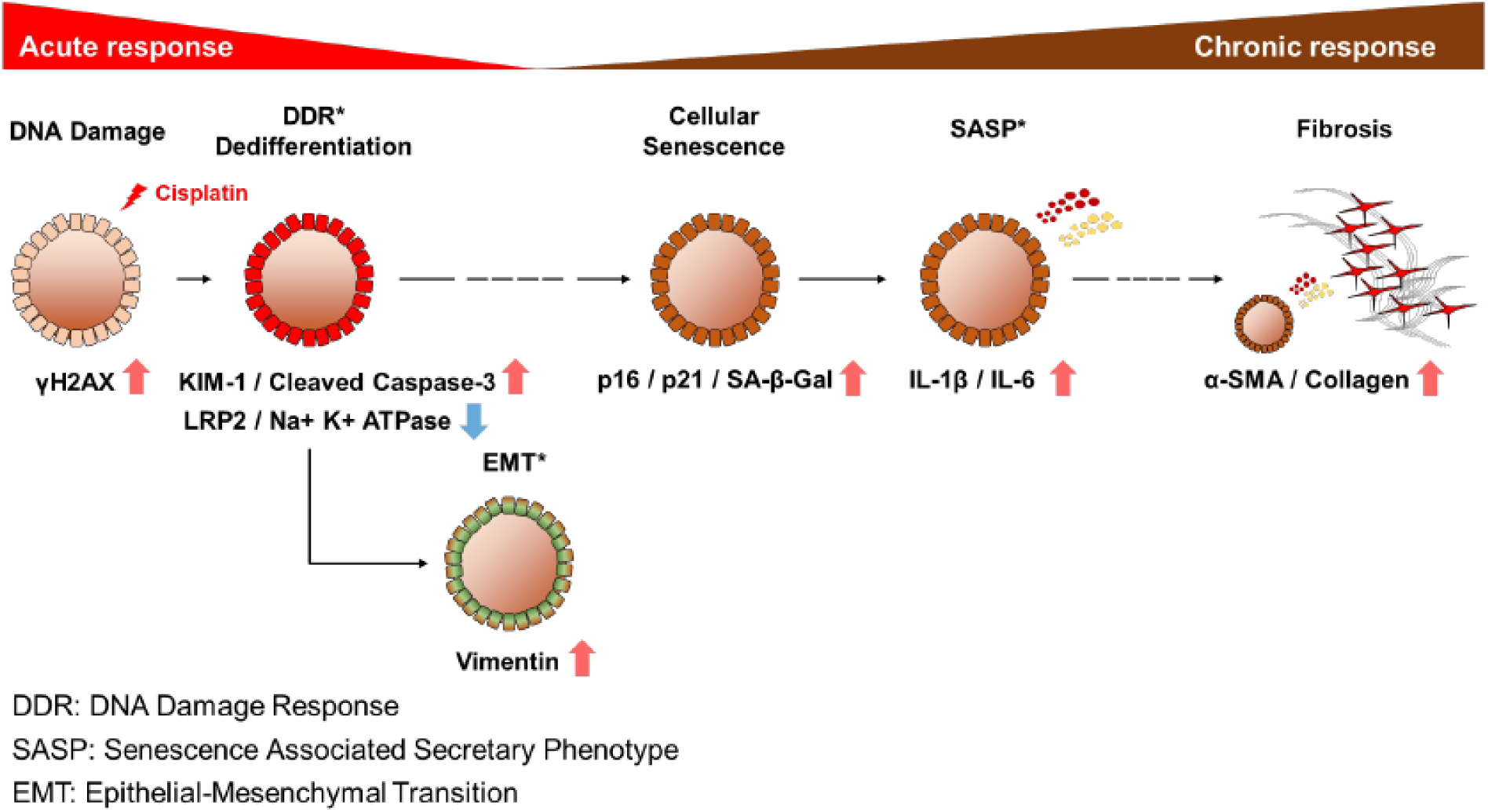

CM obtained from tubuloids repeatedly treated with 2.0 µg/mL cisplatin showed the most significant promotion of fibrosis **(Figure 4C-F).** This concentration of cisplatin, as shown in **Figure 3**, corresponds to the maximal secretion levels of IL-1β and IL-6 from tubuloids, indicating consistency. Furthermore, direct administration of cisplatin to mouse fibroblasts did not result in myofibroblast activation, decisively ruling out cisplatin as a direct fibrosis inducer on fibroblasts. The fibrosis bioassay confirmed that various paracrine factors secreted from tubuloids during repeated treatment of cisplatin have a fibrogenic effect on tissues.

## Discussion

In this study, we succeeded to recapitulate almost all the maladaptive responses of renal epithelial cells against cisplatin-induced toxicity by patient-derived tubuloids, not animal models or iPS kidney organoids. By using multiple patients-derived tubuloid, our strategy has a potential to use tubuloids as a semi-peresonalized toxicity or drug eficaciy platform. Furthermore, as we succeeded to induce senescence markers, SA-β-GAL, p16 and p21 expression, and SASP, our tubuloids can be not a drug toxicity model but also a new pathophysiological model for AKI to CKD transition or CKD model itself.

Our findings provide compelling evidence of the negative effects of cisplatin on tubuloids, elucidating the cascade of events initiated by DNA damage induction. The dose-dependent increase in γH2AX expression is a reliable indicator of DNA damage, supporting previous findings. Upon DNA damage, tubuloids exhibit a sensitive acute phase response as evidenced by increased expression of the kidney injury marker KIM-1 and the apoptosis marker Cleaved Caspase-3, which is consistent with DDR activation. There was also an increase in Vimentin expression, which indicates epithelial-mesenchymal transition (EMT).

The well-preserved three-dimensional structure of tubuloids collapsed upon exposure to cisplatin, accompanied by a process of dedifferentiation marked by reduced expression levels of Na^+^/K^+^-ATPase and LRP2/Megalin. Furthermore, repeated long-term administration of cisplatin-increased the expression of the cellular senescence markers, SA-β-GAL, p16 and p21. Tubuloids developed a senescence-associated secretory phenotype (SASP), releasing inflammatory cytokines like IL-1β and IL-6, causing a chronic inflammatory response in surrounding tissues. These secretions from tubuloids significantly increased fibrosis in the surrounding interstitial area, as confirmed by the fibrosis bioassay.

CKD research is difficult due to the complexity of the underlying conditions. A significant impediment to research into CKD is the lack of a reliable disease model. CKD is inextricably linked to aging and senescence, making it particularly difficult to model in experimental settings. Currently, the primary models for studying CKD are mice, which, despite their utility, live only about two years. This timeframe does not adequately represent the decades-long progression of chronic diseases such as CKD in humans. Furthermore, mouse models are a highly homogeneous population, lacking the diversity observed in human CKD cases. Given the variability in CKD progression among humans, which is influenced by factors such as gender, race, age, and environmental exposures, mouse models fail to capture the complexities of human CKD adequately. This discrepancy reveals a significant gap in our current understanding, emphasizing the need for more robust and clinically relevant models to advance CKD research.

In recent years, kidney organoids derived from induced pluripotent stem cells (iPSCs) or embryonic stem cells (ESCs) have emerged as promising disease models for the study of kidney diseases [30–32]. However, current technologies primarily produce organoids that look like fetal kidneys, making it difficult to replicate age-related features associated with CKD in older people [33]. Furthermore, iPSC-derived kidney organoids require complex differentiation induction procedures and skilled techniques, complicating their application. Primary cultured cells derived from resected human kidneys and kept in two-dimensional conditions fail to fully replicate the cellular polarity and three-dimensional structures found in living tissues. As a result, while these models provide valuable insights, they have limitations that prevent them from accurately representing the complexity of CKD pathogenesis and progression.

Conventional disease models for CKD, such as mouse models, iPSC/ESC kidney organoids, and two-dimensional primary cultured cells from human kidneys, struggle to fully replicate aging, senescence, and ease of production. However, the human kidney-derived tubuloids developed in our study provide significant improvements over traditional pathological models for CKD. Tubuloids are formed by cells obtained from patients that reflect their age and may exhibit senescence, as evidenced by the control condition without cisplatin treatment **(Figure 3B, C, E and G)**. This unique feature addresses the limitation of other models by incorporating the aging process directly into the system.

The formation of tubuloids follows a remarkably simple protocol, requiring no specialized techniques. Culture is carried out in standard 24-well ultra-low attachement plates, and as shown in **Figure 1F**, multiple tubuloids develop within each well. During sample collection for western blotting, qPCR, and immunofluorescence, we harvested one sample per well as an independent condition, resulting in multiple tubuloids in each sample, effectively reducing variability across individual tubuloids.

Expanding our study to include tubuloids from multiple patients shows promise for capturing individual differences in kidney responses to various stimuli. Notably, our tubuloid production protocol allows us to derive these structures from different patients simultaneously, under uniform conditions (manuscript in preparation). These characteristics point to tubuloids’ potential as a novel and useful screening tool for renal toxicity in drug discovery efforts.

Tubuloids have a highly differentiated three-dimensional structure[34] and mimic physiological responses to nephrotoxicants[35, 36]. Previous research has shown that these well-differentiated tubuloids express a diverse set of channels and transporters, providing them with greater physiological functionality than cells cultured in two dimensions [37]. This increased physiological fidelity is likely to explain the observed sensitivity to cisplatin in our study, facilitating the replication of cellular senescence and other key responses. Importantly, tubuloids may capture biological reactions that are difficult to detect in conventional two-dimensional cell cultures.

The primary focus of this study was cisplatin-induced nephropathy. However, our overarching goal goes beyond this single study to develop a strong pathophysiological model for CKD using human kidney-derived tubuloids. The precise mechanisms underlying CKD following cisplatin chemotherapy remain largely unknown. However, even low-dose cisplatin has been shown to cause long-term renal pathologies similar to CKD [38]. Cisplatin-induced nephropathy is caused by the uptake of cisplatin into proximal tubular epithelial cells via OCT2, resulting in direct damage to nuclear DNA. This DNA damage initiates a cascade of DNA damage response (DDR) processes: some cells die via programmed cell death, such as apoptosis, while others activate the cell cycle in an attempt to compensate for the loss of damaged cells. When DNA damage reaches a critical threshold, it causes cell cycle arrest, particularly in the G2/M phase[2], preventing further division, a sign of cellular senescence.

Senescent cells have been shown to activate the SASP, resulting in the secretion of a variety of inflammatory and fibrotic cytokines. This cascade causes inflammation and fibrosis in the surrounding tissue while remaining resistant to cell death. This mechanism, which is driven by cellular senescence and SASP induced by DNA damage, is a significant contributor to renal fibrosis. It is an important part of the transition from AKI to CKD, as well as the broader pathophysiology of CKD, which goes beyond cisplatin-induced nephropathy.

In our tubuloid model, we found not only a highly differentiated three-dimensional structure, but also pathophysiological responses to cisplatin, such as DDR, structural collapse, dedifferentiation, cell death, epithelial-mesenchymal transition (EMT), cellular senescence, SASP activation, and fibrotic changes. Our findings suggest that tubuloids have the potential to serve as a precise disease model for cisplatin-induced nephropathy, faithfully replicating the majority of pathological mechanisms. This underscores their commitment to developing a comprehensive CKD model.

Moving forward, we plan to improve the sophistication of our disease models by incorporating coculture systems and microfluidic technologies. Specifically, we intend to coculture tubuloids with other relevant cell types, including vascular endothelial cells and fibroblasts. Additionally, we plan to simulate physiological urine flow using microfluidic devices to better mimic the *in vivo* renal microenvironment [39].

The successful development of a highly precise disease model for cisplatin-induced nephropathy using human kidney-derived tubuloids represents a significant advance in kidney disease research. Using tubuloids as a reliable kidney disease model has great potential for improving our understanding of various kidney diseases. With their ability to faithfully replicate cellular senescence, SASP, and fibrosis, tubuloids appear to be a promising tool for modeling CKD, a condition defined by fibrosis as a final common pathological pathway.

In conclusion, our development of tubuloids as a precise disease model for cisplatin-induced nephropathy is a significant step forward in kidney disease research, especially as a presice pathophyisiological model based on patient-derived materials, not animal models or iPSC kidney organoids. Through continued refinement and innovation, we anticipate that tubuloids will play an important role in sensing drug toxicity in a semi-personalized fashion and advancing our understanding of CKD.

## Acknowledgement

We would like to thank the study participants who kindly allowed to give us the pieces of their resected kidneys.

## Data availability statements

Further information and requests for resources and reagents should be directed to and will be fulfilled by the Lead Contact, Yutaro Mori.

## Author contributions

Y.N., M.M., Y.S., I.O., R.S. and Y.M. performed the experiments, collected and analyzed data, and wrote the manuscript. Y.M., M.M., Y.S., I.O., and R.S. established the hRPTECs and S.M. helped the procedure. T.F., H.K., F.A., K.S., T.M., E.S., and S.U. supported the data analysis. Y.W., S.Y. and Y.F. resected the patients’ kidneys as standard treatment for malignant diseases. Y.M. developed experimental strategy, supervised the project, and edited the manuscript. All authors discussed the results and implications and commented on the manuscript.

